# Determinants of delayed antenatal visit attendance in rural Burkina Faso: a cross-sectional study

**DOI:** 10.1101/2025.10.22.25338535

**Authors:** Joël D. Bognini, Toussaint Rouamba, Helen Brotherton, Guétawendé J. W. Nassa, Athanase M. Somé, Diagniagou Lankoandé, Edmond Yabré Sawadogo, Umberto D’Alessandro, Halidou Tinto, Anna Roca

## Abstract

**Introduction:** The World Health Organization recommends a minimum of eight antenatal care (ANC) contacts, with the first visit occurring before the 12^th^ week of gestation, as a strategy to enhance the preparedness of women for institutional delivery and improve perinatal outcomes. The present study aims to assess the prevalence of delayed ANC attendance among pregnant women in rural Burkina Faso and identify associated risk factors.

**Methods:** This is a secondary analysis of clinical data collected from a randomised-controlled trial (clinicaltrials.gov ref: NCT03199547); conducted between 2018 and 2021 in rural Burkina Faso. We estimated gestational age (GA) at the first ANC visit based on recall information on the last menstrual period provided by study participants or, when such information was unavailable, symphysis-fundal height measurements taken by ANC nurses. We used descriptive methods followed by unadjusted and adjusted logistic regression, informed by an original conceptual framework, to determine the prevalence and risk factors associated with delayed first ANC visit, defined as occurring after the 12^th^ week of gestation. A significance threshold was set at 0.05.

**Results:** Out of the 5250 women enrolled in the study, 2480 (47.2%) had data available from their first ANC visit, and 90.6% (2248/2480) of those women had gestational age estimates. Most women (n=2037/2248, 90.6%) attended their first ANC after the 14^th^ week of gestation. The main factors associated with this delay were multiparity ≥ 4 pregnancies (OR=2.26, 95%CI [1.48 – 3.4], p < 0.001) and first ANC visit attended during the dry season (OR=1.79, 95%CI [1.34 – 2.39], p < 0.001).

**Conclusion:** Our study highlights that most pregnant women in rural Burkina Faso attended their first ANC visit later than the WHO recommended timeline, increasing their risk of poor delivery outcome. Although we identified some factors that increased this risk of late ANC attendance, awareness raising interventions are required for the whole population as starting late seems to be the norm.

## Introduction

The antenatal and peripartum periods carry significant risks of morbidity and mortality for both mothers and their newborns. In 2020, the global maternal mortality rate (MMR) was 216 deaths per 100,000 live births [1]. In sub-Saharan Africa (SSA), the MMR was substantially higher, at 536 maternal deaths per 100,000 live births [2]. Furthermore, the neonatal mortality rate (NMR) in SSA was estimated at 27 deaths per 1,000 live births in 2021 and this accounted for 45% of global neonatal deaths [3].

Professional antenatal care (ANC) is crucial for improving pregnancy outcomes. The World Health Organization (WHO) recommends a minimum of eight ANC contacts, with the first visit occurring before 12 weeks of gestation or during the first trimester of pregnancy [4]. Early attendance and regular ANC follow-ups play a crucial role in preventing and managing maternal health problems and preparing women for institutional delivery [5–7]. Indeed, primary risk factors for perinatal mortality can be identified during pregnancy, and the utilisation of at least one early ANC visit by a skilled provider reduces the neonatal mortality rate by 39% in SSA [7, 8]. A systematic review of 74 studies published between 2008 and 2018 across 23 countries in SSA, including West African nations (Burkina Faso, Nigeria, Benin, Ghana, and Niger), revealed that more than two-thirds of pregnant women attended ANC late, and up to 1 in 14 had their first visit during the third trimester of pregnancy [9].

Late ANC attendance is associated with perinatal complications and adverse birth outcomes such as increased risk of stillbirth, low birth weight, neonatal mortality and preterm delivery [5, 10–12]. These serious outcomes are important public health challenges in Burkina Faso, with 31 preterm births per 1000 live births, and 22 stillbirths for 1000 total births in 2020 [13]. In Burkina Faso, data from the Ministry of Health showed that the proportion of pregnant women who attended ANC late varied across geographic areas, ranging from 52% to 74% in 2018 [14] and approximately 60% in 2020 [13]. Despite the implementation of free health care policy in Burkina Faso in 2016, which aimed to improve access to health care services coverage and timing of ANC visits [15], there is no significant improvement of the situation [13]. To reduce the perinatal mortality rate even with the free policy, it is necessary to assess the determinants of late ANC attendance in order to be able to implement strategies to improve earlier ANC attendance in line with the WHO recommendations that the first prenatal consultation should be carried out during the first trimester (before 12 weeks of gestation) of pregnancy in an effort to reduce perinatal mortality. Hence, it is important to determine the prevalence of delayed ANC attendance and assess its main determinants in rural Burkina Faso, in the context of a free healthcare policy for pregnant women and lactating mothers.

## Methods

### Study design

This is a secondary data analysis of a descriptive cohort from the PregnAnZI-2 project (clinicaltrials.gov ref: NCT03199547); the latter is a phase III, double-blind, placebo-controlled randomised clinical trial conducted in The Gambia and Burkina Faso from 2017 to 2021. The trial procedures were reported elsewhere [16]. Briefly, women were approached for consent during ANC visits but were enrolled into the trial only during labour when oral intrapartum azithromycin or placebo was administered, hence the allocation did not influence the attendance or timing of ANC and both trial arms were included in this study. However, we have included here only the Burkina Faso cohort as it is more representative of a rural West African population compared to peri-urban Gambian participants.

### Study site

Burkina Faso is a West African country with approximately 20 million inhabitants, where the main livelihood is agriculture. There are two seasons: a dry season (November to May) and a rainy season (June to October). Women of childbearing age represent 24.1% of the population, with a fertility rate in 2020 of 5 children per woman [17, 18]. At the national level, in 2020 nearly three-quarters of pregnant women attended one ANC visit, and among them, one-quarter attended during the first trimester of pregnancy [13]. Regarding adverse perinatal outcomes in Burkina Faso, the stillbirth rate was 22 per 1000 live births, NMR of 64 per 1000 live births, and MMR of 147 per 100,000 live births in 2020 [13]. Since 2016, all services offered at health facilities are free of charge for pregnant women, lactating mothers, and children under 5 years. The present study was conducted in the Nanoro Health District catchment area located at approximately 85 km from Ouagadougou, the capital of Burkina Faso. This is a rural area with ANC services provided by midwives at government primary health centres.

### Participants

This study included pregnant women aged 16 and above recruited in the PregnANZI-2 trial and who delivered at the study health facility with ANC services available. Women were excluded if: they had a known acute or chronic condition (e.g. HIV infection, diabetes mellitus); had a planned caesarean section or anticipated referral to a tertiary referral facility; known severe foetal congenital malformation; macrolide allergy or use of medication known to increase the QT interval during two weeks preceding delivery [16].

### Study procedures

Written informed consent for participation in the PregnAnZI-2 trial was taken during ANC visits and verbally confirmed during labour. Socio-demographic, epidemiological, and clinical information were collected by trained research nurses during labour and the immediate post-partum period (typically 6-24h after delivery) using the handheld Maternal and Child Health book and maternal direct questioning, as per PregnAnZI-2 trial procedures [16].

### Outcome measures and variables of interest

The outcome of interest is a binary variable; delayed ANC attendance (yes or no) defined as first visit after 14^th^ week of gestation. This variable was generated using the gestational age (GA) recorded during the first ANC visit to classify weeks of pregnancy. We estimated GA at the first ANC visit based on recall information of the last menstrual period (LMP) provided by study women or when such information was not recalled by the women, we used symphysis-fundal height measurements assessed by the government facility nurses. The correlation between fundal height and the GA in weeks was previously reported to have a prediction error ranging between 13.9 and 14.9 days [19]. Gold standard dating methods using first trimester ultrasound scanning were not available either as part of routine antenatal care or in association with PregnAnZI-2 trial.

To explore factors associated with delayed ANC attendance, we developed an original conceptual framework based on the literature review and plausible associations (Figure 1). Although existing frameworks exist for measuring ANC quality of care [20] and for ANC in well-resourced settings [21], we did not identify any conceptual frameworks relevant to our population of West African women living in a rural low-resource setting. Hence, we developed an original one. The conceptual framework guided our choice of variables for the univariate, unadjusted analyses and informed the development of adjusted analytical models. The categories for these variables were selected based on a preliminary analysis of the data to consider small cell sizes, and similar outcomes in adjacent ordered or related categories.

**Figure 1.**
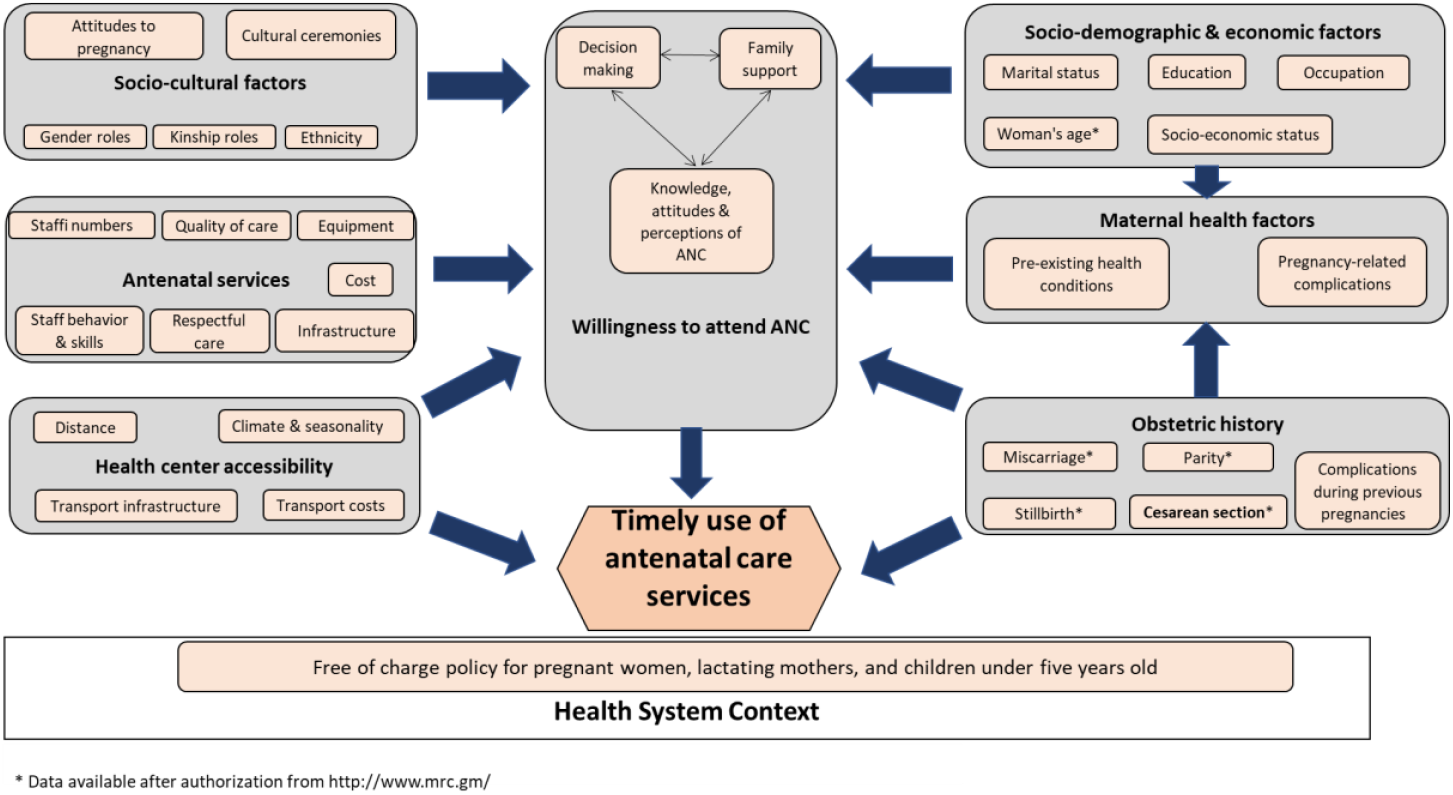
Conceptual framework to understand determinants of pregnant women’s timely use of antenatal care services

### Data management and statistical analyses

All data were electronically captured using the REDCap software, version 8.9.2, with internal validation and consistency checks. Descriptive statistics (percentage, mean, and standard deviations) were performed to determine the prevalence of all independent variables. Logistic regression was used to assess the association between each covariate and to generate crude odds ratio and 95% confidence intervals (CI). Variables with p-value < 0.20 from the unadjusted analyses that were part of the conceptual framework were included in the adjusted models. A p-value < 0.05 was considered as statistically significant. All analyses were conducted using Stata 15.

## Results

### Characteristics of the study population

Overall, 42.8% (2248/5250) of enrolled women had available information regarding the timing of their first ANC visit and were included in the analysis. Out of them, 1% recalled their LMP and in the remaining 99% fundal height measurements were taken by a study nurse. Demographic and epidemiological characteristics of women included, and those who were not included in the study were not statistically significantly different, except that the included women were slightly younger (Supplementary Table 1). For included women, mean age was 26.5 years, and more than half were multiparous. Around 58% of pregnant women attended their first ANC during the dry season (November-May), 9% and 3% had history of miscarriage and stillbirth respectively (Table 1). Overall, 2037/2248 (90.6%) of women attended their first ANC visit after 14 weeks of gestation. The majority of these pregnant women (75.7%, 1701/2248) had their first ANC visit during the second trimester, and 14.9% (336/2248) during the third trimester of their pregnancy (Figure 2).

**Table 1:** Characteristics associated with delayed ANC attendance in rural Burkina Faso.

|  | Total | Delayed ANC visit attendance |  | Unadjusted analysis |  |  | Adjusted analysis |  |  |
| --- | --- | --- | --- | --- | --- | --- | --- | --- | --- |
| Characteristics | Total<br>2248 (100%) | No<br>211 (9.4%) | Yes<br>2037 (90.6%) | OR | CI 95% | p-value | OR | CI 95% | p-value |
| **Age of the pregnant woman (Years)** |  |  |  |  |  | 0.01* |  |  | 0.28 |
| **Age (mean, SD)** | 26.51 (6.33) | 24.49 (5.69) | 26.72 (6.35) |  |  |  |  |  |  |
| 20 – 35 | 1,680 (74.73) | 159 (9.46) | 1521 (90.54) | 1 |  |  | 1 |  |  |
| 15 – 19 | 362 (16.11) | 43 (11.88) | 319 (88.12) | 0.77 | [0.54 -1.10] |  | 1.33 | [0.81 – 2.20] |  |
| > 35 | 206 (9.16) | 09 (4.37) | 197 (95.63) | 2.28 | [1.15 – 4.55] |  | 1.49 | [0.73 – 3.05] |  |
| **Mothers' ethnicity** |  |  |  |  |  | 0.37 |  |  |  |
| Mossi | 2,077 (92.39) | 197 (9.48) | 1,880 (90.52) | 1 |  |  |  |  |  |
| Gurunsi | 144 (6.41) | 10 (6.94) | 134 (93.06) | 1.40 | [0.72 – 2.71] |  |  |  |  |
| Others | 27 (1.20) | 4 (14.81) | 23 (85.19) | 0.60 | [0.20 – 1.75] |  |  |  |  |
| **Husbands' ethnicity** |  |  |  |  |  | 0.55 |  |  |  |
| Mossi | 2,070 (92.08) | 197 (09.52) | 1,897 (90.48) | 1 |  |  |  |  |  |
| Gurunsi | 143 (6.36) | 10 (06.99) | 133 (93.01) | 1.39 | [0.72 – 2.70] |  |  |  |  |
| Others | 35 (1.56) | 4 (11.43) | 31 (88.57) | 0.81 | [0.28 – 2.33] |  |  |  |  |
| **Period of the ANC visit commencement** |  |  |  |  |  | < 0.001* |  |  | < 0.001 |
| Rainy (June-October) | 947 (42.13) | 116 (12.25) | 831 (87.75) | 1 |  |  | 1 |  |  |
| Dry (November-May) | 1,301 (57.87) | 95 (7.30) | 1,206 (92.70) | 1.77 | [1.33 – 2.35] |  | 1.79 | [1.34 – 2.39] |  |
| **Parity** |  |  |  |  |  | < 0.001* |  |  | < 0.001 |
| Para 2 | 347 (15.44) | 43 (12.39) | 204 (87.61) | 1 |  |  | 1 |  |  |
| Primiparous | 438 (19.48) | 59 (13.47) | 379 (86.53) | 0.90 | [0.59 – 1.38] |  | 0.77 | [0.46 – 1.27] |  |
| Para 3 | 335 (14.90) | 44 (13.13) | 291 (86.87) | 0.93 | [0.59 – 1.46] |  | 0.93 | [0.59 – 1.47] |  |
| Para ≥ 4 | 1,128 (50.18) | 65 (5.76) | 1,063 (94.24) | 2.31 | [1.54 – 3.47] |  | 2.26 | [1.48 – 3.45] |  |
| **History of miscarriage** |  |  |  |  |  | 0.23 |  |  |  |
| No | 2,043 (90.88) | 187 (9.15) | 1,856 (90.85) | 1 |  |  |  |  |  |
| Yes | 205 (9.12) | 24 (11.71) | 181 (88.29) | 0.65 | [0.59 – 4.58] |  |  |  |  |
| **History of stillbirth** |  |  |  |  |  | 0.33 |  |  |  |
| No | 2,181 (97.02) | 207 (9.49) | 1,974 (90.51) | 1 |  |  |  |  |  |
| Yes | 67 (2.98) | 4 (5.97) | 64 (94.03) | 1.96 | [0.61 – 6.31] |  |  |  |  |

\* Variables included in the adjusted model.
Abbreviations: ANC = Antenatal care; CI = Confidence interval; OR = Odds ratio; SD = Standard deviation

**Figure 2:**
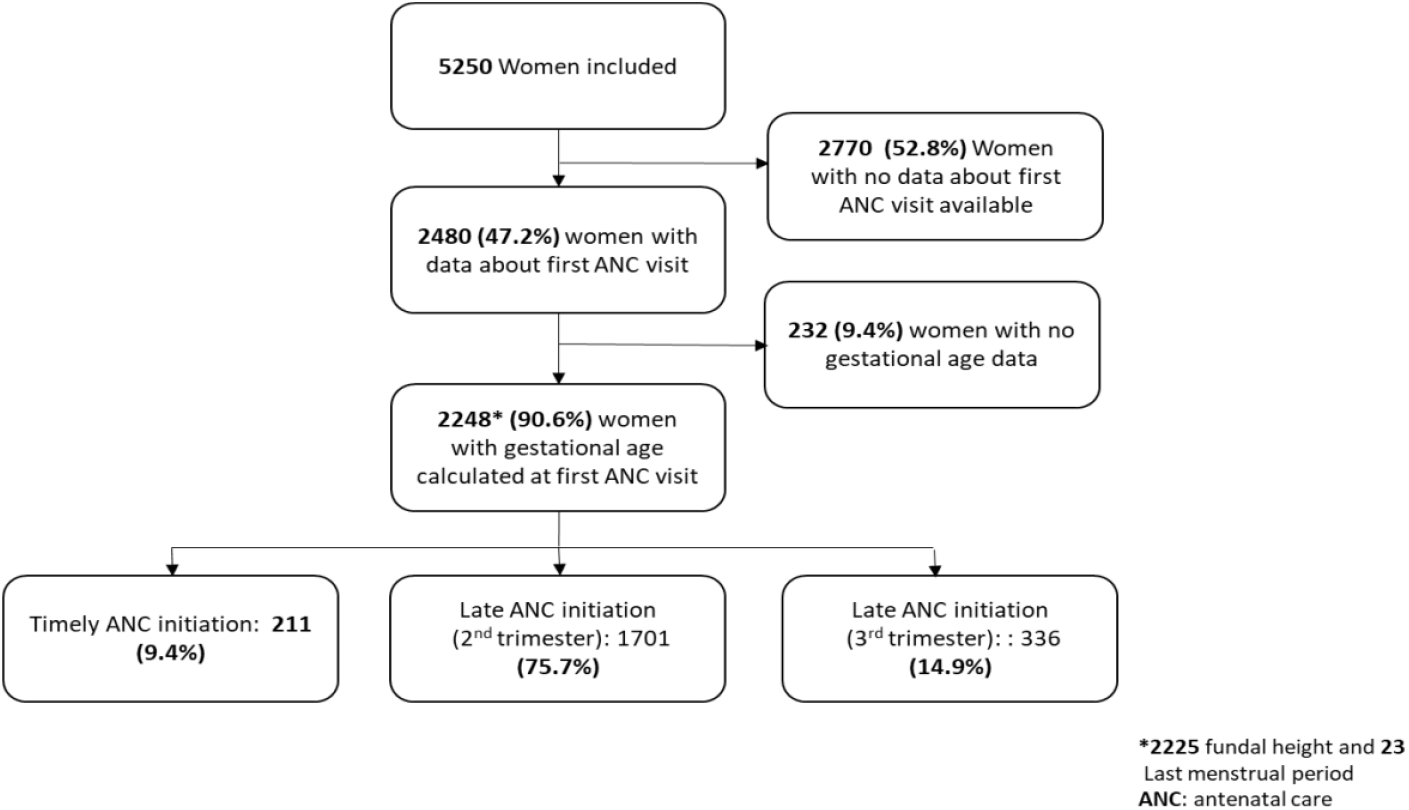
Flowchart showing the inclusion of relatively healthy pregnant women in the study.

### Determinants of delayed ANC attendance

Parity and season of the first ANC visit were associated with delayed ANC attendance according to the adjusted regression model. Multiparous women pregnant for fourth or more time had an increased odds of starting ANC visits at > 14 weeks of gestation compared to women in their second pregnancy (OR=2.26 95%CI [1.48 – 3.35], p < 0.001). Women who attended their first ANC visit during the dry season were more likely to attend after 14 weeks of gestation compared to those attending for the first time during the rainy season (OR=1.79, 95%CI [1.34 – 2.39], p < 0.001) (Table 1). Even though, we found statistically significant differences for the prevalence of delayed ANC attendance between parity groups, less than 15% of women within each parity group had their first ANC visit on time; and the same applies to both seasons.

## Discussion

This large cohort study highlights that most pregnant women in rural Burkina Faso do not attend their first ANC visit before the 14^th^ week of gestation, even after the implementation of a free national healthcare policy. This delay occurs across all age groups, ethnic groups, parities and seasons.

In our study, at least 90% of relatively healthy pregnant women living in rural Burkina Faso attended their ANC visits after the 14^th^ week of gestation, later than the WHO recommended timeline. This figure exceeds the delayed ANC attendance previously reported from Burkina Faso at national level (62.2% to 64.3%) [22]. Similar rates of late ANC attendance were reported at another health district level (62.9%) located in the Centre-North of Burkina Faso [23]. Prevalence of delayed ANC attendance also varies between countries across West Africa, ranging from 85.3% in Nigeria [24] to 57.7% in Ghana [25]. Elsewhere in Africa, delayed ANC attendance rates are also a concern as reported in rural Ethiopia, Eastern Africa (60.5%) [23, 26, 27], albeit not as high as observed in our study.

Reasons for the high prevalence of delayed ANC attendance in our rural West African study population are likely multifactorial. Potential factors include local beliefs that revealing pregnancy too early would result in miscarriage, financial constraints for transport to health facilities, and limitation in the availability and quality of the ANC services. These issues are not unique to Burkina Faso as they have been observed also in South Africa, Tanzania and other African countries [28–30], as depicted in the framework. This framework could facilitate the identification of factors for delay at first ANC visit in similar contexts for other studies. The widespread delay in ANC attendance poses significant risks as it exposes pregnant women to adverse pregnancy outcomes [31] by missing early preventive care and the timely identification of maternal, foetal and obstetric complications. To address this, emphasising the importance of early ANC visits both at health facility level and in the communities through primary health workers could enhance awareness and promote earlier engagement with ANC services, as demonstrated in Ethiopia [32] and Uganda [33], where pregnant women with knowledge of appropriate ANC timings were more likely to initiate care timely. In Burkina Faso, despite free ANC services, economic barriers such as the cost of specialised tests, transportation and the indirect economic impact of time spent away from formal or domestic work still pose a significant challenge. Innovative evidence-based solutions are needed to overcome these barriers and ensure more women can access timely to ANC. Also, introducing a more flexible ANC schedule system could further improve the early attendance.

Although most women were late to their first ANC visit, we identified two important determinants: multiparity and season of first ANC visit. Consistent with previous studies from Ethiopia [32] and Nigeria [34], multiparous women in our study were more than twice as likely to have delayed ANC attendance compared to women during their second pregnancy. A possible explanation, supported by a qualitative study in Cameroon, is that women with extensive previous experience of pregnancy gain confidence in their own ability to manage their pregnancies and perceive early healthcare as less critical [35]. An alternative explanation could be that attitudes were shaped by the women’s previous negative experiences in the health facility, such as long waiting times and poor quality of care, as highlighted in a study from Papua New Guinea [36].

We observed an association between season of first ANC visit and timely attendance, with increased risk of delay during the dry season. This finding may be linked to domestic work demands for pregnant women, as reported in an Ethiopian study where the time constraints linked to household activities represented more than 24% of reasons for delayed ANC attendance [27]. During the dry season in rural Burkina Faso there are various cultural activities which coincide with the harvest campaign, including mask dances and funeral ceremonies [37–39], hence women are busy with domestic duties and this may impact on the attendance of their first ANC visit. Even though we identified two determinants of delayed first ANC visit, most women were at risk of being late and therefore any intervention to improve early ANC attendance should ideally target all of them.

This study has some limitations that need to be considered when interpreting the results. Firstly, we used data from a selected population participating in a randomised controlled trial, which could have introduced potential bias by excluding women with serious health problems or those not willing to participate. Secondly, gestational age was estimated using inaccurate measures (last menstrual period and symphysis-fundal height), limiting the accuracy of the estimations and potentially misclassifying the primary outcome. However, we mitigated this risk by using an established method with precision of 2 weeks to minimise the misclassification errors. A third limitation is the selection bias towards pregnant women with available data to estimate the gestational age. However, comparison of baseline characteristics for included and excluded pregnant women did not identify any major differences in their characteristics, hence bias is as assumed to be minimal. Lastly, as this analysis used an existing dataset, we were unable to assess all potential variables associated with delayed ANC attendance. Hence, there is a need for further qualitative and quantitative research to understand determinants of timely attendance of ANC for rural West African populations.

## Conclusion

Our study identified a very high proportion of pregnant women in rural Burkina Faso attending their ANC visits later than the WHO recommended timeframe with multiparity and dry season the main determinants of delayed ANC initiation. This delay poses significant public health concerns by limiting the opportunities for maternal and antepartum preventive services. Merely providing free access to ANC or implementing national policies has proven to be insufficient in ensuring women starting their ANC on time. There is a pressing need for further research to understand the context-specific reasons behind these delays to optimise maternal and perinatal health outcomes effectively.

## Data Availability

Data may be obtained from a third party and are not publicly available. Qualified researchers may request access with the Gambia Government/MRC Joint Ethics Committee. The review process and release of data will be facilitated by MRC Unit The Gambia (http://www.mrc.gm/) through the Head of Governance at MRCG. Access will not be unduly restricted.

## DECLARATIONS

### Ethics approval and consent to participate

The trial was approved by the ‘‘Comité d’Ethique pour la Recherche en Santé (CERS)’’ and the Ministry of Health of Burkina Faso, The Gambia Government/MRCG (Medical Research Council Unit The Gambia) Joint Ethics Committee and the LSHTM Ethics Committee. All women provided written informed consent during antenatal care visits and were free to withdraw at any time.

### Consent to publish

Not applicable.

### Competing interests

The authors declare that they have no competing interests.

### Funding

The PregnAnZI-2 trial was funded by a grant from the UKRI under the Joint Global Health Trial Scheme (JGHT) (ref: MC_EX_MR/P006949/1) and the Gates Foundation (Ref: OPP1196513). The funders and study sponsor (MRCG) had no role in the study design, collection, analysis, or interpretation of data, writing of the article nor the decision to submit for publication.

### Authors’ contributions

J.D.B. conceptualised this study with input from U.dA., H.B., H.T., A.R. Data collection was coordinated by J.D.B., and PregnAnZI-2 field teams in Burkina Faso. J.D.B. performed the analysis with full access to the data and input from T.R. J.D.B. drafted the initial manuscript with input from H.B., A.R., H.T., T.R., A.M.S., G.J.W.N., E.Y.S, D.L. All authors contributed to the final version. H.T. and A.R. gave oversight to the work as guarantors and accepted full responsibility for the finished work and controlled the decision to publish.

## Acknowledgements

The authors wish to thank Medical Research Council Unit, The Gambia at LSHTM for allowing access to the data, the research staff, and all the participants of Nanoro health district for accepting to take part in the study.

## Notes

### Competing Interest Statement

The authors have declared no competing interest.

### Clinical Trial

NCT03199547

### Author Declarations

The parent trial and use of data was approved by the Comite d'Ethique pour la Recherche en Sante and the Ministry of Health of Burkina Faso, the Gambia Government/MRCG (Medical Research Council Unit The Gambia) Joint Ethics Committee and the LSHTM Ethics Committee. All women provided written informed consent during antenatal care visits and were free to withdraw at any time.

