## Supplementary Table 1 for "Determinants of delayed antenatal visit attendance in rural Burkina Faso: a cross-sectional study"

Supplementary Table 1. Comparison of characteristics between women who were included and excluded from the study

| **Characteristics** | **Excluded** | **Included** | **p-value** |
| --- | --- | --- | --- |
|  | **N=3002**  **n (%)** | **N=2248**  **n (%)** |  |
| **Age (years); mean (SD)** | 26.1 (6.3) | 26.5 (6.3) | 0.05 |
| **Age (years); median (Q25 - Q75)** | 26 (21 – 30) | 26 (21 – 30) |  |
| **Age of the pregnant woman (Years)** |  |  | 0.01 |
| 15 – 25 | 1499 (49.93) | 1041 (46.31) |  |
| 26 – 35 | 1270 (42.31) | 1001 (44.5) |  |
| > 35 | 233 (7.76) | 206 (9.16) |  |
| **History of abortion** |  |  | 0.38 |
| Yes | 295 (9.83) | 205 (9.12) |  |
| No | 2705 (90.17) | 2043 (90.88) |  |
| **History of stillbirth** |  |  | 0.39 |
| Yes | 102 (3.40) | 67 (2.98) |  |
| No | 2898 (96.60) | 2181 (97.02) |  |
| **Caesarean section** |  |  | 0.12 |
| Yes | 6 (0.2) | 1 (0.04) |  |
| No | 2994 (99.8) | 2247 (99.96) |  |
| **Mother ethnicity** |  |  | 0.74 |
| Mossi | 2759 (91.91) | 2077 (92.39) |  |
| Gurunsi | 201 (6.7) | 144 (6.41) |  |
| Others | 42 (1.4) | 27 (1.2) |  |
| **Husband ethnicity** |  |  | 0.79 |
| Mossi | 2759 (91.94) | 2070 (92.08) |  |
| Gurunsi | 188 (6.26) | 143 (6.36) |  |
| Others | 54 (1.8) | 35 (1.56) |  |
| **Period of the antenatal visit commencement** |  |  | 0.63 |
| November-May (Dry) | 138 (59.48) | 1301 (57.87) |  |
| June-October (Rainy) | 94 (40.52) | 947 (42.13) |  |
| **Parity** |  |  | 0.44 |
| Primiparous | 612 (20.4) | 438 (19.48) |  |
| Secondi-parous | 487 (16.23) | 347 (15.44) |  |
| Multiparity | 1901 (63.37) | 1463 (65.08) |  |
